# Statistical Evaluation of the Test Threshold for the Alzheimer’s Disease EEG Coherence Marker

**DOI:** 10.1101/2022.09.07.22279698

**Authors:** Crystal Radinski, Grace Perez

## Abstract

A progressive reduction in synaptic connections between neurons is one neurophysiological indicator of brain ageing and was linked to the severity of dementia. In our study, we hypothesized that if synaptic disconnection as the neuropathology of Alzheimer’s disease (AD) is responsible for the brain’s inability to integrate diverse regions into efficient networks, then electroencephalographic data may be utilized to identify Alzheimer’s dementia. The study explored EEG coherence, reflecting connectivity between regions, as a potential AD indicator and identified four promising EEG coherence markers. As the pattern of degeneration follows the temporal-parietal-frontal axis, the temporal gamma marker was chosen for further evaluation. We utilize conventional analysis, producing paired results such as sensitivity and specificity and the receiver operating characteristic (ROC) curve to evaluate the accuracy of the temporal gamma marker. The optimal cut-off point of 0.950 was confirmed by both methods and provided 95% correct classification indicating an almost perfect differentiation between control and impaired cognitive status. This evaluation will be used in a blinded diagnostic test accuracy study to determine the TG_marker validity in detecting AD and excluding pseudo-dementias.

---

A progressive reduction in synaptic plasticity and synaptic connections between neurons is one neurophysiological indicator of brain ageing and was linked to the severity of dementia [1]. In our study, we hypothesized that if synaptic disconnection as the neuropathology of Alzheimer’s disease (AD) is responsible for the brain’s inability to integrate diverse regions into efficient networks, then electroencephalographic data may be utilized to identify Alzheimer’s dementia [2]. The study explored EEG coherence, reflecting connectivity between regions, as a potential AD marker. We examined group differences in EEG coherence within global cortical networks at rest and during executive challenges among patients with AD, individuals with mild cognitive impairment, and healthy controls. Task-related EEG coherence for cross-hemisphere electrode pairs in four brain regions (frontal F3-4, parietal P7-8, temporal T7-8, occipital O1-2) was evaluated with MATLAB software [3,4]. A statistically significant decrease in EEG coherence (p<0.05) has been discovered in cross-hemisphere frontal, temporal, parietal and occipital pairs in the AD group at rest and when challenged with tasks requiring comprehension, analysis, perceptual-motor response, and executive functioning. The four most promising EEG coherence markers were identified as (i) F3-F4 Beta during visual-spatial orientation task (p=0.019), P7-P8 Beta during writing (p=0.001), (iii) T7-T8 Gamma during multistep command challenge (p=0.008) and (iv) O1-O2 Alpha during, space orientation task (p=0.020).

Medial temporal lobe atrophy and decreased hippocampus volume are the most typical focused MRI findings in AD [5]. The pattern of degeneration follows the temporal-parietal-frontal axis [6]. Although neuronal degeneration in AD is a diffuse prosses, the earliest cortical neuronal disconnection seems to be most noticeable in the temporal cortical region. Therefore, the T7-T8 Gamma marker was chosen for further evaluation (TG_marker).

## I. Descriptive Statistics

Out of 14 tests in neurocognitive testing, the multistep command challenge was particularly effective in revealing neuronal disconnection in temporal lobes. The test consists of a verbal three-step command requiring a participant to “take the paper in your right hand, fold it and place it on the table.” A participant must listen to the entire three-step direction before proceeding and executing the steps in the order they were listed. The three-step command is a common test in verified neurocognitive test panels like Mini-Mental State Examination [7]. The task recruits left superior temporal and inferior parietal regions. The descriptive statistics and test results for TG_marker coherence measures for each group across the 10 EEG runs during the test are summarized in Table 1.

**Table 1.**
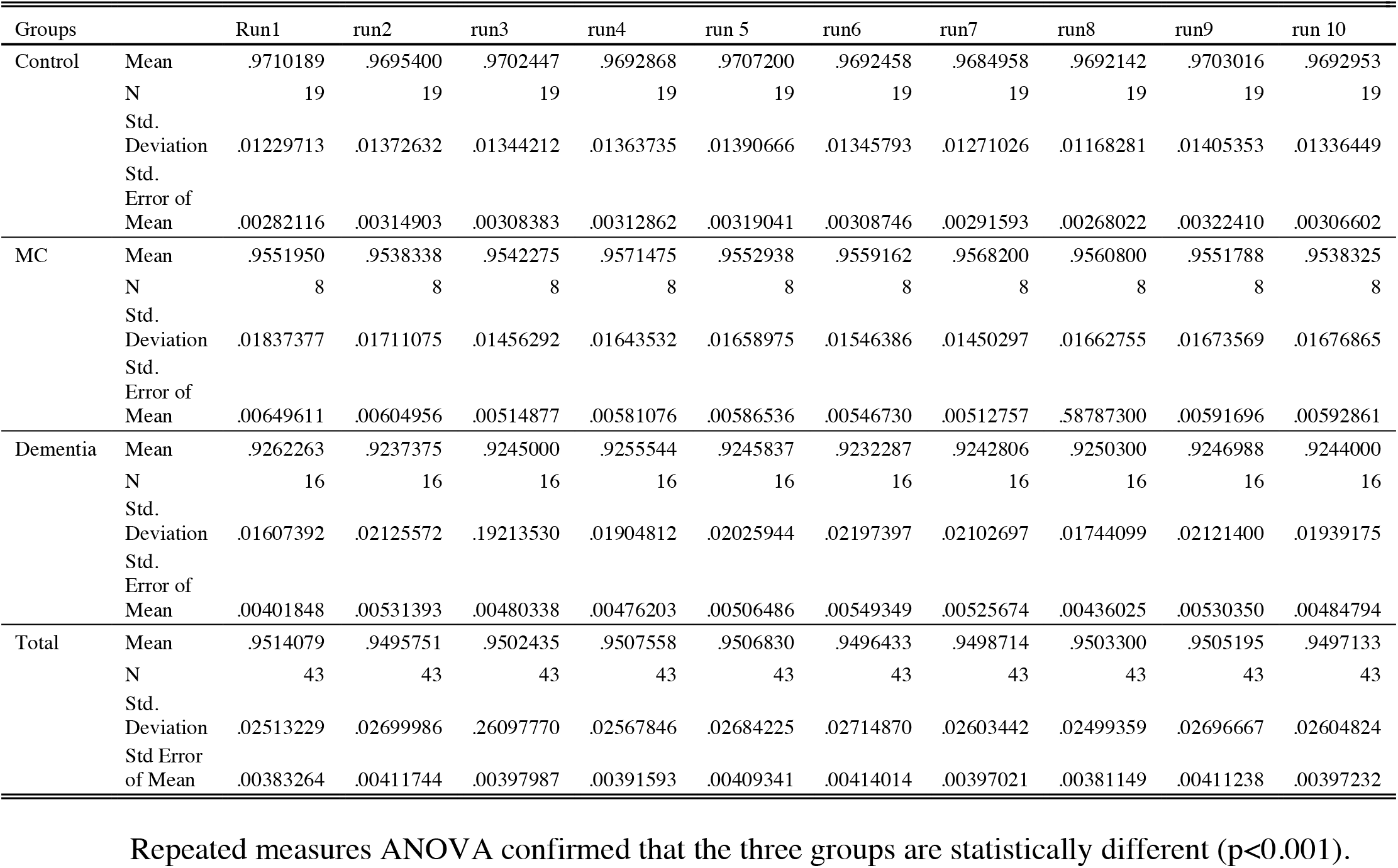
TG_marker data for the control, dementia and MCI groups, across 10 EEG runs in three-step command test.

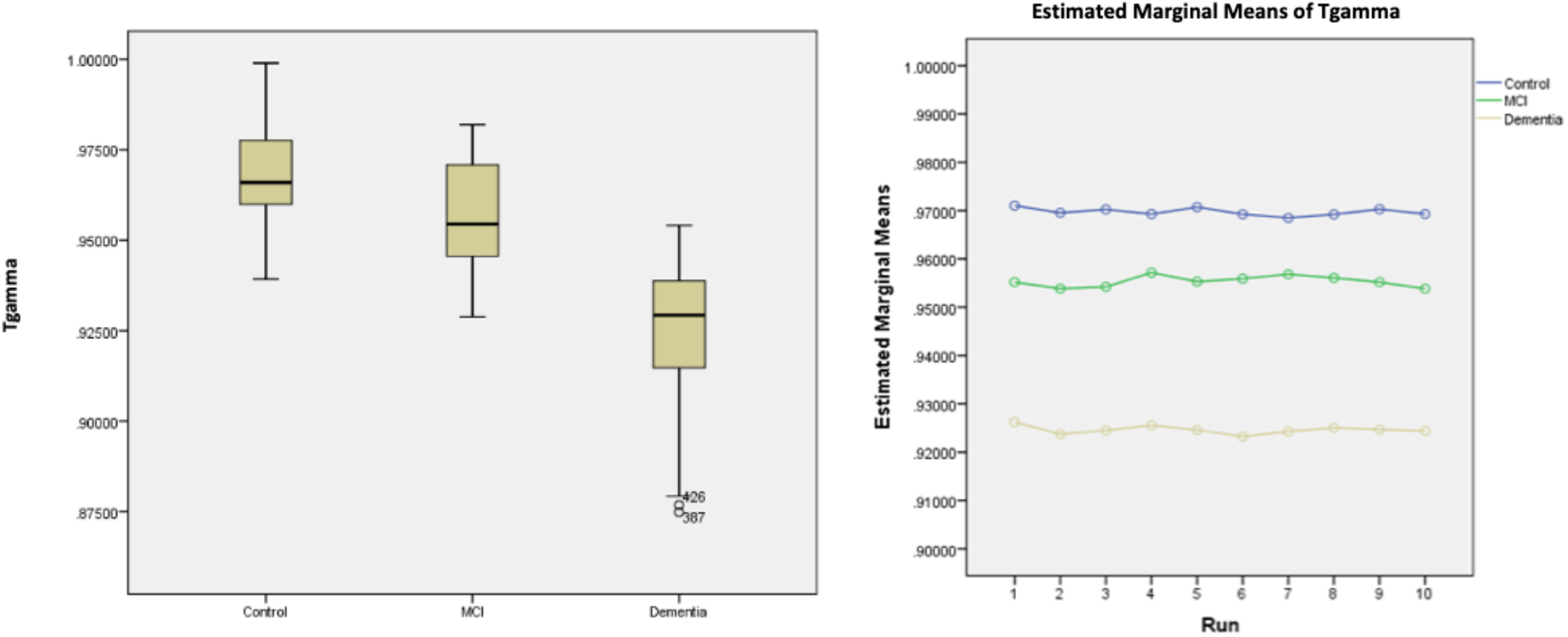

## II. TG_Marker as Diagnostic Test for Dementia

### 2.1 Determining Cut-off Points for the Marker as Diagnostic Test for Dementia

EEG coherence represents the functional interaction between two regions [8, 9]. It is an advantageous method for exploring the functioning of neuronal networks. EEG coherence ranges from 0 to 1, with higher values indicating full task coherence while lower values indicating poor task coherence. Poor task coherence may indicate neuronal disconnection resulting in cognitive function decline. AD group demonstrated significantly lower EEG coherence than the control group in temporal lobes. (p<0.001).

In order to find potential cut-off points, we analyzed the distribution of the TG_marker values for AD and the control group. We analyzed the data from nineteen control participants and sixteen AD group members. As the test was segmented in 10 epochs, there were190 data points available for the control and 160 data points for the dementia group (Table 2).

**Table 2.** TG_marker data for the control and dementia groups

| Groups | Mean | N | Std. Deviation | Std. Error of Mean | Minimum | Maximum |
| --- | --- | --- | --- | --- | --- | --- |
| Control | .9697363 | 190 | .01295029 | .00093951 | .93922 | .99889 |
| Dementia | .9246240 | 160 | .01921776 | .00151930 | .87486 | .95407 |
| Total | .9491135 | 350 | .02766895 | .00147897 | .87486 | .99889 |

The distribution of the temporal gamma values for the control and dementia groups within ±2SD of the mean, which contains at least 95% of the values, is demonstrated in the two graphs below.

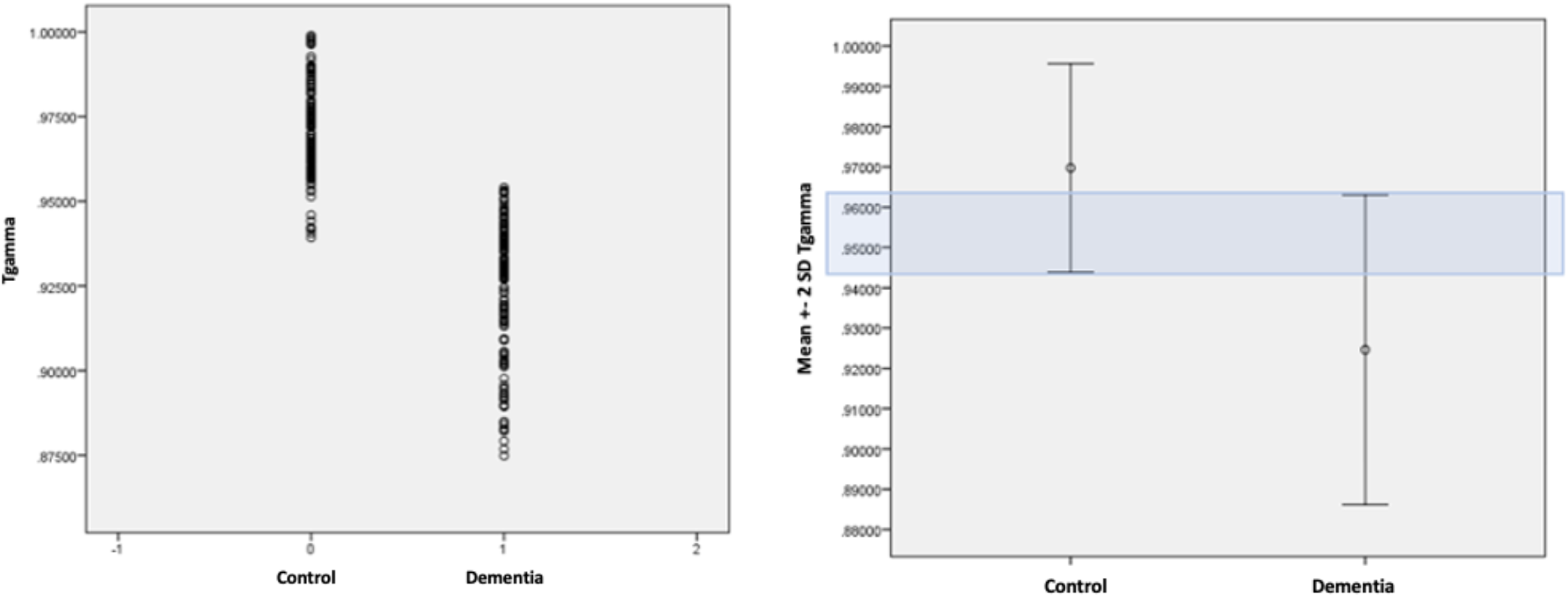

The distribution of the temporal gamma values for the control and dementia groups within ±2SD of the mean.

The marker distributions for the control and AD groups intersect above 0.940 up to below 0.965.

### 2.2 Evaluation of Cut-off Points as Diagnostic Test for Dementia

Exploratory study: To evaluate the accuracy of the TG_marker as a diagnostic test for AD, we followed the conventional way of describing diagnostic test outcomes (positive/negative results) when compared with the gold standard, as demonstrated in table 3. The gold standard, in this case, is the actual clinical diagnosis of the true disease state for dementia. Diagnostic accuracy can be presented at a specific threshold by using paired results such as sensitivity and specificity, positive predictive value (PPV) and negative predictive value (NPV).

**Table 3.**
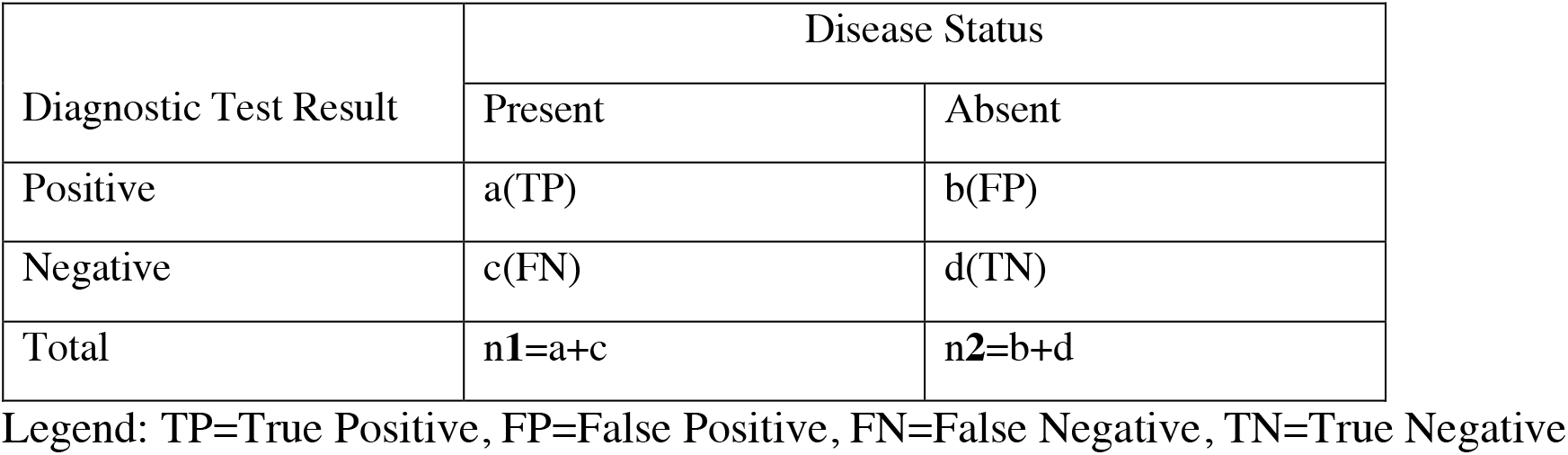
Summary Indices of Test Performance

Conventional analyses consider the sensitivity and specificity of a diagnostic test as the primary indices of accuracy since these indices are independent of the prior probability of disease.

|  |  |
| --- | --- |
| Sensitivity = $TP/(TP+FN) = a/(a+c)$ | Positive predictive value = $TP/(TP+FP) = a/(a+b)$ |
| Specificity = $TN/(FP+TN) = d/(b+d)$ | Negative predictive value = $TN/(FN+TN) = d/(c+d)$ |

For tests that produce data on a continuous scale, testing thresholds must be specified in order to distinguish between positive and negative findings. The percentage of false positive and false negative diagnoses varies when the threshold is altered. We analyzed several cut-off points in multiples of 5 thousandth points (0.940, 0.945, 0.950, 0.955, 0.960 and 9.965) covering an intersecting area of the control and AD groups distributions above 0.940 up to below 0.965 (Table 4).

**Table 4.**
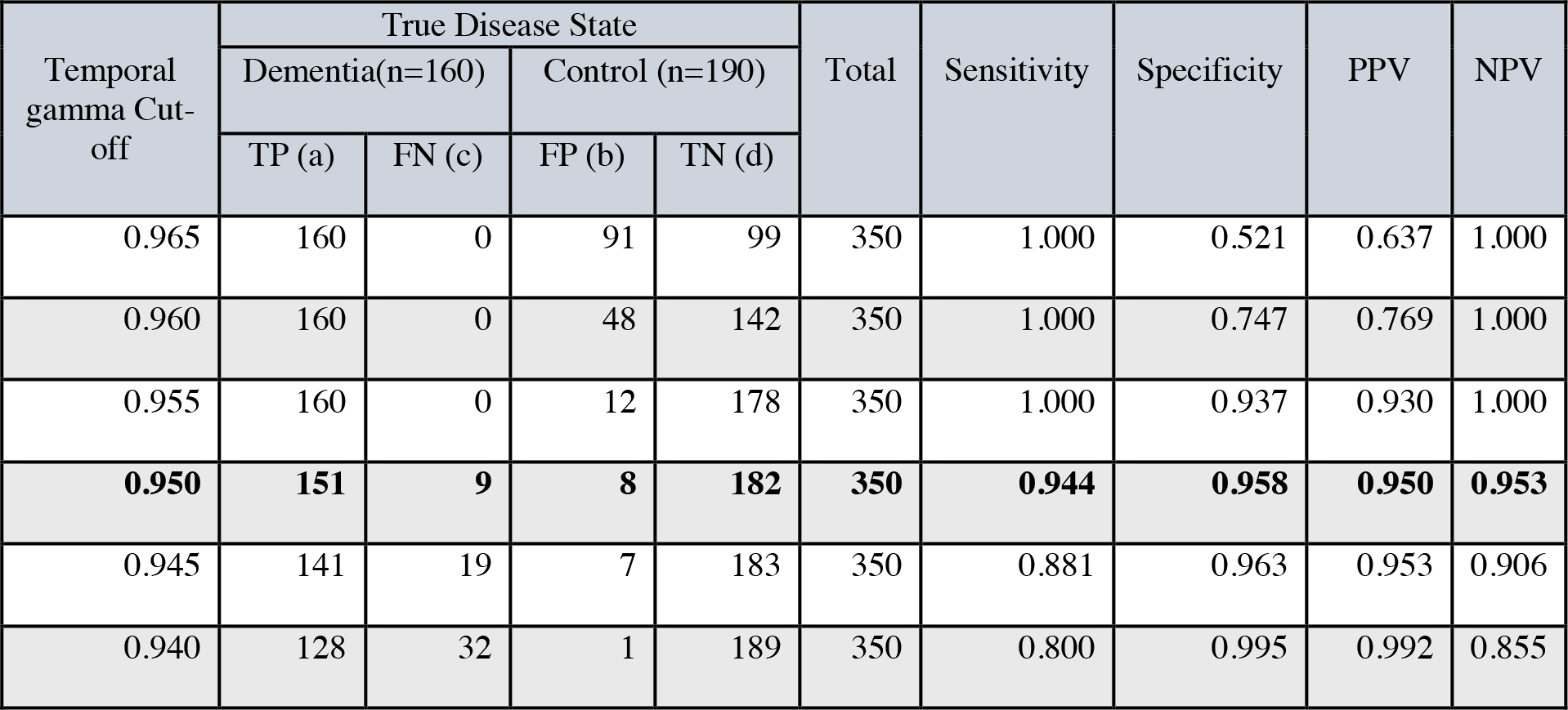
Sensitivity, specificity, PPV and NPV at multiple cut-off points of TG-marker

As demonstrated in Table 4, the cut-off point of 0.950 appears to be of the optimal value, providing the best balance between sensitivity (94.4%) and specificity (95.8%).

### 2.3 ROC Curve Analysis for TG_Marker as Diagnostic Test

The application of the receiver operating characteristic (ROC) curve is another way to evaluate the accuracy of the TG_marker as a diagnostic test for AD, where diagnostic accuracy is summarised by combining across a range of thresholds [10]. A ROC curve is the graphical representation of the reciprocal relationship between sensitivity and specificity and shows a single test accuracy at different thresholds. The ROC curve is important in determining how well diagnostic tests can distinguish between subjects’ real states. The analysis has been used extensively in clinical epidemiology for the assessment of the diagnostic ability of markers and imaging tests in the classification of diseased from healthy subjects [11,12].

Logistic regression analysis was utilized to examine TG_marker values as a predictor of AD (1=AD and 0=control or no AD) in order to compute the probability estimates of the predicted group classification. The classification table produced by logistic regression demonstrated that the TG_marker correctly classified 95% of the cases. The manually created distribution table matches the cut-off point of 0.950 in the first evaluation demonstrating the same result.

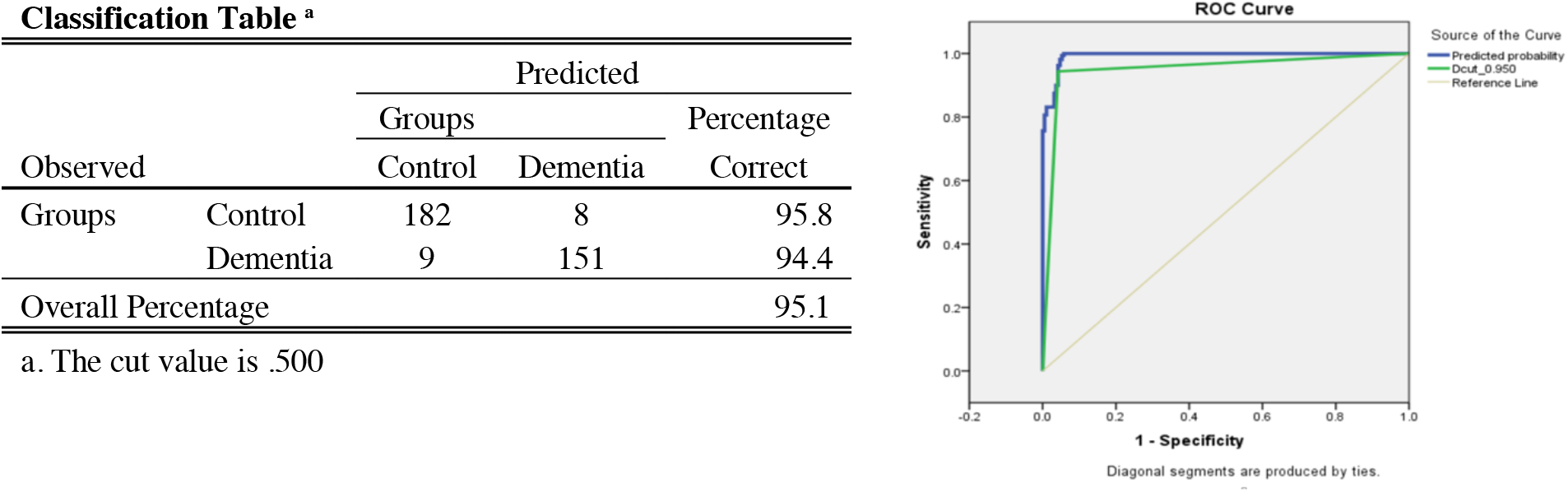

The ROC curves for both the actual and grouped temporal gamma values are shown in blue and green, respectively. The diagonal line is the reference line for the area-under-the-curve (AUC), which is set by default at 0.50.

**Area Under the Curve**
| Test Result Variable(s) | Area | std. Error<sup>a</sup> | Asymptotic Sig<sup>b</sup> | Asymptotic 95% Confidence Interval |  |
| --- | --- | --- | --- | --- | --- |
|  |  |  |  | Lower Bound | Upper Bound |
| Predicted probability | .993 | .003 | .000 | .987 | .998 |
| Dcut_0.950 | .951 | .013 | .000 | .924 | .977 |

The test result variable(s): Dcut\_0.950 has at least one tie between the positive actual state group and the negative actual state group. Statistics may be biased.
a. Under the nonparametric assumption
b. Null hypothesis: true area = 0.5

The area under the curve for TG_marker values is 0.993, with p<0.001. The logistic regression model classified the group significantly better than mere chance alone.

## III. Conclusion for TG_Marker as Diagnostic Test

Logistic regression and ROC curve analyses confirmed the classification table that resulted in the optimal cut-off point of 0.950. This cut-off point provided 95% correct classification, and the corresponding area under the curve 99.3%, indicating an almost perfect differentiation between control and impaired cognitive status.

No diagnostic test has perfect accuracy, and all tests occasionally fail to detect disease or perceive it in healthy patients. However, false negative and false positive diagnoses carry unequal significance. The misclassification cost, the relative importance of a false negative versus a false positive diagnosis, varies according to the disease’s effect on patients and the effectiveness of available treatments. Timely detection of a life-threatening disease for which a cure is available and time-sensitive is more important than a false positive diagnosis in a healthy patient. In the case of AD, the false positive diagnosis can trigger immense anxiety in patients and their caregivers and increase the cost to the healthcare system with further investigations. However, the false negative will not cause patients to forgo the benefit of disease-modifying treatment. Recognizing reversible causes of neurocognitive impairment could be even more critical as curative or quality-of-life-improving treatments could be available for pseud-dementias such as those caused by mood disorders and metabolic abnormalities.

Evaluation of the TG_marker in a blinded diagnostic test accuracy study is needed to determine the test validity in detecting AD and excluding pseudo-dementias.

## Data Availability

All data produced in the present work are contained in the manuscript

